# Pulmonary Embolism in Patients with COVID-19: A Systematic review and Meta-analysis

**DOI:** 10.1101/2020.10.09.20209965

**Authors:** Omar Hamam, Ahmed Goda, Radwa Awad, Amr Ussama, Moustafa Eldalal, Ahmed Fayez, Karim Elyamany, Renu Bhandari, Waleed Ikram, Abdelrhman Elbaz, Smarika Baral, Yomna Elbandrawy, Alexander Egbe, Iraida Sharina

## Abstract

**Background:** There is an increasing evidence that COVID-19 could be complicated by coagulopathy which may lead to death; especially in severe cases. Hence, this study aimed to build concrete evidence regarding the incidence and mortality of pulmonary embolism (PE) in patients with COVID-19.

**Methods:** We performed a systematic search for trusted databases/search engines including PubMed, Scopus, Cochrane library and Web of Science. After screening, the relevant data were extracted and the incidences and mortality rates from the different included studies were pooled for meta-analysis.

**Results:** Twenty studies were finally included in our study consisting of 1896 patients. The results of the meta-analysis for the all included studies showed that the incidence of PE in patients with COVID-19 was 17.6% with the 95% confidence interval (CI) of 12.7 to 22.5%. There was significant heterogeneity (*I*^2^□=□91.17%). Additionally, the results of meta-analysis including 8 studies showed that the mortality in patients with both PE and COVID-19 was 43.1% with the 95% confidence interval (CI) of 19 to 67.1%. There was significant heterogeneity (*I*^2^□=□86.96%).

**Conclusion:** PE was highly frequent in patients with COVID-19. The mortality in patients with both COVID-19 and PE was remarkable representing almost half of the patients. Appropriate prophylaxis and management are vital for better outcomes.

## INTRODUCTION

Coronavirus disease 2019 (COVID-19); caused by the severe acute respiratory syndrome coronavirus 2 (SARS-CoV-2) is currently a global health pandemic that threatens the lives of millions everywhere^1^. COVID-19 manifestations vary from mild respiratory symptoms to severe multi-organ failure and death^2,3^. Moreover, there is an increasing evidence that COVID-19 could be complicated by coagulopathy which may lead to death; especially in severe cases ^4,5^.

The viral infection, respiratory deterioration, and the use of central venous catheter may provide major risk factors for the occurring thromboembolism ^6^. Likewise, the activation of coagulation and thrombo-inflammation with local vascular damage may enhance the process^4-6^. Among the associated complications, pulmonary embolism (PE) has been reported^7^.

Studies showed different estimates for the epidemiological aspects PE with COVID-19. For instance, a study by Grillet et al ^8^showed that PE incidence detected by pulmonary CT angiography was 23%. Additionally, many studies showed incidence of venous thrombosis reaching up to 80% even with thromboprophylaxis^9-11^. However, the exact incidence of PE in COVID-19 patients is still not clear, being based on few observational studies without large sample sizes. Therefore, this systematic review and meta-analysis is aimed to build a concrete evidence about the incidence and mortality of PE among patients with COVID-19.

## METHODS

We adhered to the Preferred Reporting Items for Systematic Reviews and Meta-Analyses (PRISMA) guidelines and Cochrane’s handbook of systematic reviews to conduct this study^12,13^.

### Literature search

We combined the following keywords and conducted our search: “COVID-19”, “SARS-CoV-2”, “pulmonary embolism” and “venous thromboembolism”. We searched PubMed, Web of Science, Scopus, and Cochrane Library for relevant articles to be included. An additional online and manual search was performed on Google Scholar and Preprint Servers to ensure adequate inclusion of all studies.

### Eligibility criteria

Results were imported into Endnote X8 (Thompson Reuter, CA, USA) for duplicates deletion. We included valid case series (>10 patients) and cohort studies including adults with COVID-19 with pulmonary embolism. Review articles, editorial, commentaries were excluded.

### Studies selection

The first author (O.H) divided other authors into two teams; each team independently performed title and abstract screening. Then, each team obtained the full-text of the included papers and performed full-text screening. Any disagreement between the two teams was resolved through consultation with the study seniors (A.E and I.S).

### Data extraction

The two teams extracted the data; one team performed extraction of selected outcomes and the other team extracted baseline data. The data were then revised in a cross-revision manner. Extracted data include author, country, year, study design, age, sex, total number of patients, and number of patients with PE, PE diagnosis, prophylactic treatment, and mortality rate from PE patients.

### Risk of bias assessment

We used the Newcastle–Ottawa scale (NOS) which is available at (https://www.ohri.ca/programs/clinical_epidemiology/oxford.asp) for assessing the risk of bias for our included studies. The possible scores of this scale range from 0 to 9. Studies with a score of seven to nine, four to six, and zero to three were classified as studies with low, moderate, and high risk of bias, respectively.

### Data synthesis and analysis

The meta-analysis of the included studies was performed using OpenMeta [Analyst] version 1.15 for conducting single-arm meta-analysis. Meta-analysis for proportions was utilized to pool the incidence and mortality of PE in the groups. Dichotomous data were calculated to obtain risk ratios along with their 95% confidence intervals (CIs). Heterogeneity among studies was assessed using the I^2^ test and P-value from the chi-squared test of heterogeneity. Values of I^2^ >50 and P<0.1 are significant markers of heterogeneity among studies according to Cochrane’s handbook^13^. Random effect models were used to avoid the effect heterogeneity. The statistical significance was set with P-value at 0.05.

## RESULTS

### Results of the literature search

We searched the aforementioned search engines/databases and found 1452 studies after duplicate removal. We excluded 1432 studies as they were not eligible for inclusion according to eligibility criteria, and a total of 20 studies were finally included in our study consisting of 1896 patients^14-33^. Figure (1) shows a summary of our search and table (1) shows the summary of the included studies.

### Baseline characteristics

Baseline characteristics are shown in table 1. Most of the included studies were conducted in Europe with only one report from USA. Among the included studies, the highest mean age was 73 years while the lowest was 60.5 years. Most of the included studies have a male predominance reaching 83% of the total included patients. CT pulmonary angiography was used for diagnosis of PE in most of the included studies.

**Table 1.** Study characteristics of the included studies.

| **First author (Year)** | **Study design** | **Country** | **Sample size** | **Male (%)** | **Age (Mean/median) years** | **PE diagnosis** | **Prophylactic treatment (%)** | **NOS** |
| --- | --- | --- | --- | --- | --- | --- | --- | --- |
| Artifoni (2020) | Retrospective cohort | France | 71 | 61 | 64 | CTPA | 99 | 7 |
| Fraissé (2020) | Retrospective cohort | France | 72 | 79 | 61 | CDU | 47 | 7 |
| Thomas (2020) | Retrospective cohort | UK | 63 | 69 | NA | CTPA | NA | 6 |
| Lodigiani (2020) | Retrospective cohort | Italy | 362 | 68 | 66 | CTPA | 79 | 8 |
| Riker (2020) | Case series | USA | 16 | NA | NA | CTPA | NA | 6 |
| Wichmann (2020) | Case series | Germany | 12 | 75 | 73 | Autopsy | 33 | 6 |
| Klok (2020) | Retrospective cohort | Netherlands | 184 | 76 | 64 | CTPA | 100 | 7 |
| Llitjos (2020) | Retrospective cohort | France | 26 | 77 | 68 | CDU | 31 | 7 |
| Helms J (2020) | Prospective cohort | France | 150 | 81 | 63 | CTPA | 100 | 7 |
| Menter (2020) | Retrospective cohort | Switzerland | 21 | 81 | 76 | Autopsy | NA | 7 |
| Florian (2020) | Retrospective cohort | France | 135 | 70 | 64 | CTPA | 53 | 7 |
| Hékimian (2020) | Retrospective cohort | France | 51 | NA | NA | CTPA or autopsy | NA | 7 |
| Longchamp (2020) | Case series | Switzerland | 25 | 64 | 68 | CTPA | 96 | 7 |
| Leonard-Lorant (2020) | Retrospective cohort | France | 106 | 66 | 64 | CTPA | 46 | 7 |
| Grillet | Retrospective | France | 51 | 70 | 66 | CTPA | NA | 7 |

### Risk of bias assessment

Among our twenty included studies evaluated for the risk of bias, all studies had a low risk of bias with score of six or higher (Table 1).

### Incidence of PE in COVID-19

The results of meta-analysis including the 20 studies showed that the incidence of PE in patients with COVID-19 was 17.6% with the 95% confidence interval (CI) of 12.7 to 22.5%. There was significant heterogeneity (*I*^2^□=□91.17%) which can be attributed to the variability of reported incidences among the different included studies (Figure 2).

**Figure 1.**
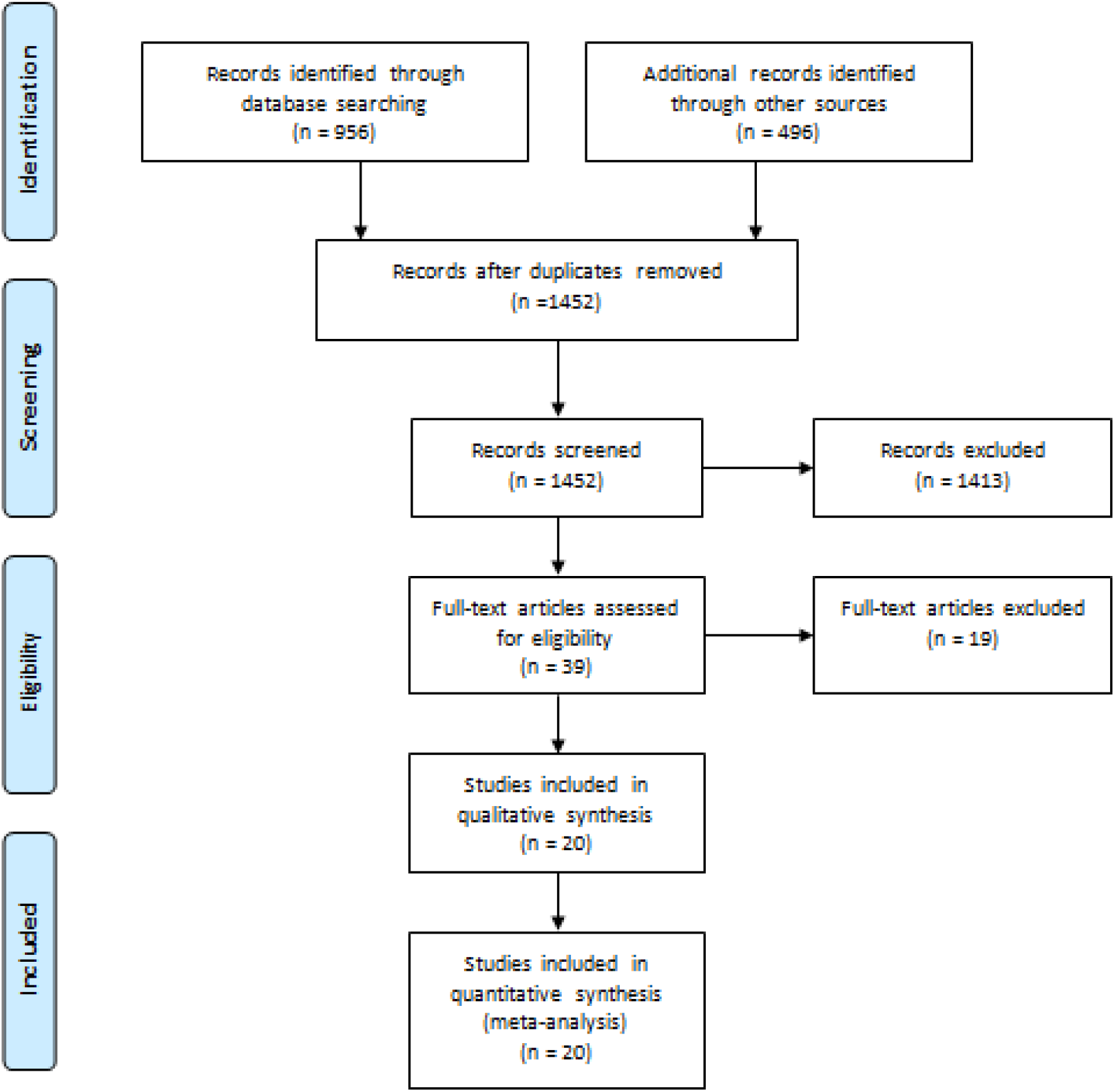
PRISMA flow diagram demonstrating the search process.

**Figure 2.**
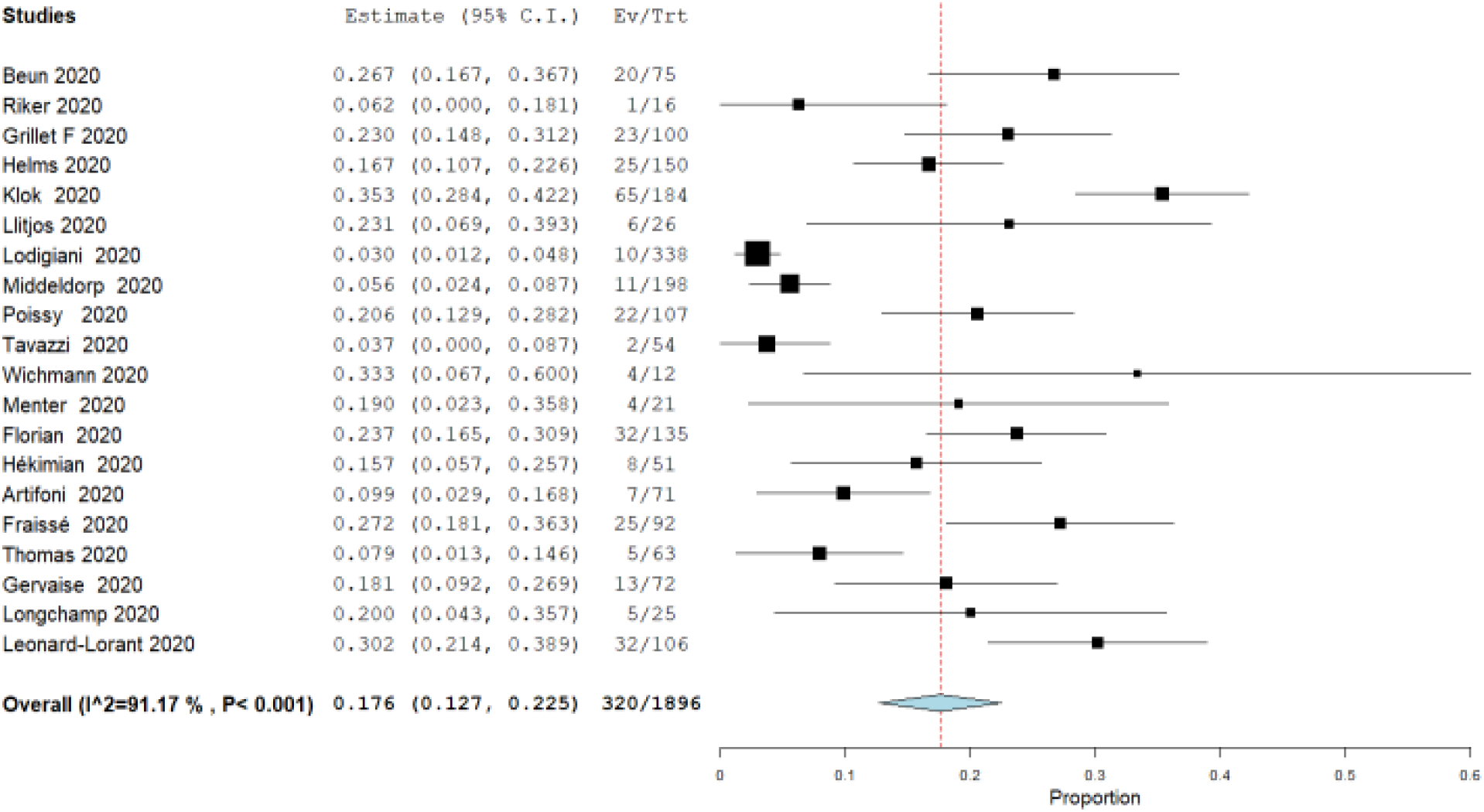
Incidence of Pulmonary embolism in COVID-19 patients.

### PE mortality in COVID-19

The results of pooled analysis including 8 studies showed that the mortality in patients with both PE and COVID-19 was 43.1% with the 95% confidence interval (CI) of 19 to 67.1%. There was significant heterogeneity (*I*^2^□=□86.96%) as shown in figure 3.

**Figure 3.**
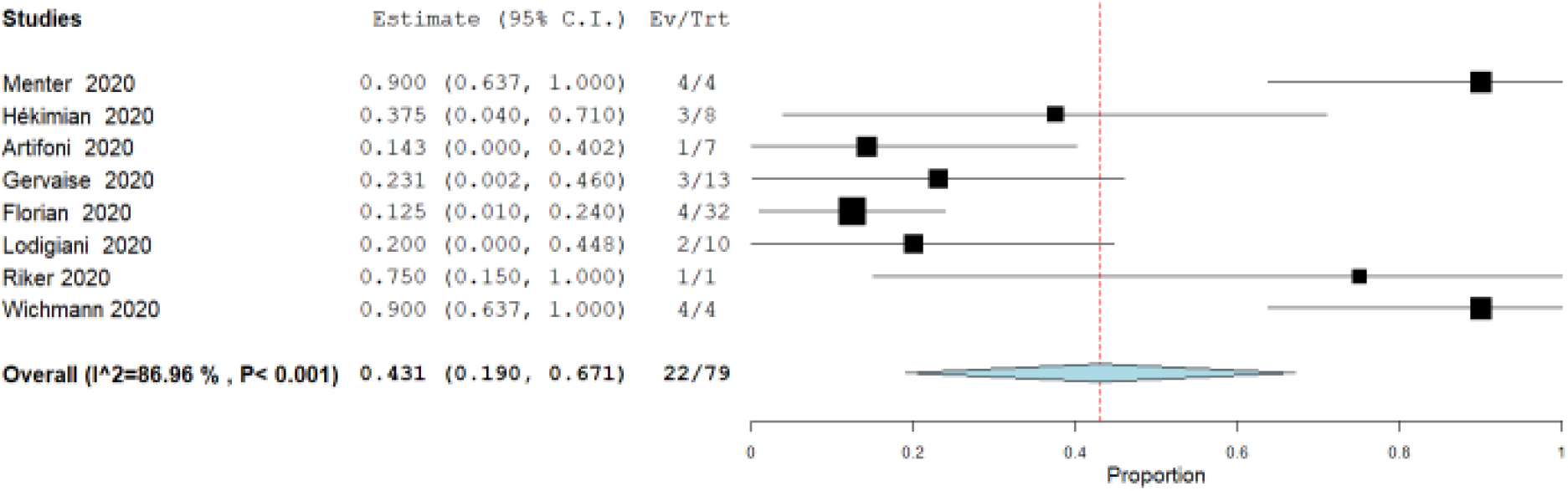
Mortality of patients with pulmonary embolism and COVID-19

## DISCUSSION

Previous studies have showed that coagulation could be a common complication among patients with COVID-19 ^5-8^. The results of this systematic review and meta-analysis showed that PE occurred frequently in patients with COVID-19 with incidence exceeding 17% of the total cases with COVID-19. This is consistent with other individual studies as in Helms et al ^15^ where the incidence of 16.7% was reported. In our study, the cumulative number of patients with both PE and COVID-19 was 320 out of 1896 patients with COVID-19, which is a slightly higher percentage than Helms et al^15^. The mortality rate in patients with both PE and COVID-19 was high, reaching 43.1% in our pooled analysis, emphasizing that PE is likely to worsen the outcome in COVID-19 patients.

The main cause of death in PE is right ventricular (RV) failure in response to RV afterload induced by thrombus mechanical obstruction of blood flow in the pulmonary arteries and pulmonary vasoconstriction. RV dysfunction is related to a short-term clinical deterioration and prognosis^34^. Reduction of the thrombus mass, a traditional approach in the PE treatment, is only partially effective and does not address the vascular humoral responses leading to increased vasoconstriction. Pulmonary vasoconstriction is an important contributor to the increase of pulmonary vascular resistance (PVR) in PE and represents an attractive therapeutic target ^35,36^. Presently, relatively small number of clinical trials or case reports exists in the literature. Inhibition of vasoconstriction by antagonists of endothelin pathway and prostaglandings were used successfully in animal models and several clinical cases to improve right ventricular function and in treatments of chronic thromboembolic pulmonary hypertension (CTEPH) ^37^. However, clinical trial on the effects of prostaglandin treatment in acute PE did not find any significant effects in comparison with placebo^38^.

Another attractive therapeutic target is manipulation of Nitric Oxide(NO)/cGMP signaling. NO/cGMP signaling plays a central role in cardiovascular system by inducing vasodilation, inhibiting leukocyte adhesion to vascular endothelium and platelet activation, among other positive hemodynamic effects ^39^. To transduce downstream signaling NO must activate the enzyme soluble guanylyl cyclase (sGC), which catalyze cGMP synthesis and represent the natural bottleneck for NO/cGMP signal transduction. Recent clinical study compared the effectiveness of inhaled NO, riociguat and cinaciguat, two new drugs directly targeting sGC, in subjects with submassive pulmonary embolism (PE) ^40^.

Riociguat, recently approved by the Food and Drug administration (FDA) for treatment of chronic thromboembolic pulmonary hypertension, is an allosteric sGC stimulator that enhances the response of NO-sensitive form of sGC ^41^. Cinaciguat, presently in phase II clinical trials by Bayer AG ^42^, preferentially activates sGC in its oxidized or heme-free states, when sGC is insensitive to both NO and nitrovasodilators. Both sGC drug exhibits potent vasodilator and antiplatelet activity and a long-lasting antihypertensive effect. In patients with acute submassive PE cinaciguat, but not riociguat or inhaled NO, effectively decreased platelets functional and metabolic hyperactivity^40^. This is likely due to persistent inflammatory conditions inducing vascular oxidative stress, which scavenges bioavailable NO and oxidizes the heme moiety of sGC, rendering the enzyme NO-insensitive. Thus, sGC direct activators, like cinaciguat, have greater pharmacological potential under pathophysiological and oxidative stress conditions compared to sGC stimulators, direct NO administration and NO donors ^43^. Application of sGC activators could be potentially useful in COVID-19 patients to manage PE.

There are different documented speculations to explain the association of PE with COVID-19. It may be correlated with viral infection, respiratory deterioration, and the use of central venous catheter that may provide major risk factors for the thromboembolism ^6,8^.

We noticed significant heterogeneity while performing this meta-analysis. This can be explained by the different and variable estimates among the included studies and different settings and severity degrees among COVID-19 patients in the different included studies. It is also worth noting that most our included studies did not separate by the severity degrees and progress which prevent us from subgrouping in our meta-analysis.

This study can be considered the most updated and comprehensive study to assess the incidence and mortality of PE with COVID-19 in a suitable number of patients. However, this study suffered from several limitations. The included studies were all observational retrospective cohort studies and case series and this type of studies has its own known limitations. Additionally, there was a wide variation among the reported items in the included studies, which leaded to limitation in pooling more of the expected common data for analysis.

To recapitulate, PE was highly frequent in patients with COVID-19 and observed in 17.6% of them. The mortality in patients with both PE and COVID-19 was remarkable reaching 43.1%. Appropriate prophylaxis and management are vital for better outcomes.

## Data Availability

Available upon request from the corresponding author

## Authors’ contribution

O.H. and A.G. conceived and designed the study. All authors acquired the data, performed the data extraction, and performed extensive research on the topic. A.E. and I.S. reviewed and performed extensive editing and supervision of the study and manuscript. All authors contributed to the writing of the manuscript. O.H. performed the statistical analysis.

## Acknowledgement

None

## REFERENCES

1. World Health Organization. Coronavirus disease 2019 (COVID-19) Situation Report e 95. 2020.

2. W.J. Guan, Z.Y. Ni, Y. Hu, W.H. Liang, C.Q. Ou, J.X. He, L. Liu, H. Shan, et al, Clinical characteristics of coronavirus disease 2019 in China, N. Engl. J. Med. (2020), https://doi.org/10.1056/NEJMoa2002032.

3. Wang D, Hu B, Hu C, Zhu F, Liu X, Zhang J, et al. Clinical characteristics of 138 hospitalized patients with 2019 novel coronavirus-infected pneumonia in Wuhan, China. JAMA 2020;323(11):1061. https://doi.org/10.1001/jama.2020.1585.

4. Kollias A, Kyriakoulis KG, Dimakakos E, Poulakou G, Stergiou GS, Syrigos K. Thromboembolic risk and anticoagulant therapy in COVID-19 patients: emerging evidence and call for action. Br J Haematol. (2020) 189:846–7. doi: 10.1111/bjh.16727

5. Wang T, Chen R, Liu C, Liang W, Guan W, Tang R, et al. Attention should be paid to venous thromboembolism prophylaxis in the management of COVID-19. Lancet Haematol. (2020) 7:e362–3. doi: 10.1016/S2352-3026(20)30109-5

6. Klok FA, Kruip M, van der Meer NJM, Arbous MS, Gommers D, Kant KM, et al. Incidence of thrombotic complications in critically ill ICU patients with COVID-19. Thromb Res. (2020) 191:145–7. doi: 10.1016/j.thromres.2020.04.013

7. Susen S, Tacquard CA, Godon A, Mansour A, Garrigue D, Nguyen P, et al. Prevention of thrombotic risk in hospitalized patients with COVID-19 and hemostasis monitoring. Crit Care. 2020;24(1):364.

8. Grillet F, Behr J, Calame P, Aubry S. Acute pulmonary embolism associated with COVID-19 pneumonia detected by pulmonary CT Angiography. Radiology. (2020). doi: 10.1148/radiol.2020201544.

9. N. Potere, E. Valeriani, M. Candeloro, M. Tana, E. Porreca, A. Abbate, S. Spoto, A.W.S. Rutjes, M. Di Nisio, Acute complications and mortality in hospitalized patients with coronavirus disease 2019: a systematic review and meta-analysis, Crit. Care 24 (2020) 389, https://doi.org/10.1186/s13054-020-03022-1.

10. Chen, X. Li, S. Liu, F. Wang, Prevalence of venous thromboembolism in patients with severe novel coronavirus pneumonia, J. Thromb. Haemost. (2020), https://doi.org/10.1111/jth.14830.

11. J.F. Llitjos, M. Leclerc, C. Chochois, J.M. Monsallier, M. Ramakers, M. Auvray, K. Merouani, High incidence of venous thromboembolic events in anticoagulated severe COVID-19 patients, J. Thromb. Haemost. (2020), https://doi.org/10.1111/jth.14869.

12. Moher D, Liberati A, Tetzlaff J, Altman DG, Altman D, Antes G, et al. Preferred reporting items for systematic reviews and meta-analyses: The PRISMA statement. PLoS Medicine. 2009.

13. Higgins JPT, Green S. Cochrane Handbook for Systematic Reviews of Interventions: Cochrane Book Series. Vol. Version 5. 2008. 1 p.

14. Riker RR, May TL, Fraser GL, Gagnon DJ, Bandara M, Zemrak WR, et al. Heparin-induced Thrombocytopenia with Thrombosis in COVID-19 Adult Respiratory Distress Syndrome. Res Pract Thromb Haemost. 2020;4:936–941.

15. Helms J, Tacquard C, Severac F, Leonard-Lorant I, Ohana M, Delabranche X, et al. High risk of thrombosis in patients with severe SARS-CoV-2 infection: a multicenter prospective cohort study. Intensive Care Med. 2020;46(6):1089–1098.

16. Wichmann D, Sperhake JP, Lütgehetmann M, Steurer S, Edler C, Heinemann A, et al. Autopsy Findings and Venous Thromboembolism in Patients With COVID-19. Ann Intern Med. 2020;M20–2003.

17. Klok FA, Kruip MJHA, van der Meer NJM, Arbous MS, Gommers D, Kant KM, et al. Confirmation of the high cumulative incidence of thrombotic complications in critically ill ICU patients with COVID-19: An updated analysis. Thromb Res. 2020;191:148–150.

18. Llitjos JF, Leclerc M, Chochois C, Monsallier JM, Ramakers M, Auvray M, et al. High incidence of venous thromboembolic events in anticoagulated severe COVID-19 patients. J Thromb Haemost. 2020;18(7):1743–1746.

19. Menter T, Haslbauer JD, Nienhold R, Savic S, Deigendesch H, Frank S, et al. Postmortem examination of COVID-19 patients reveals diffuse alveolar damage with severe capillary congestion and variegated findings in lungs and other organs suggesting vascular dysfunction. Histopathology. 2020; https://doi.org/10.1111/his.14134.

20. Bompard F, Monnier H, Saab I, Tordjman M, Abdoul H, Fournier L, et al. Pulmonary embolism in patients with Covid-19 pneumonia. Eur Respir J. 2020;2001365.

21. Hékimian G, Lebreton G, Bréchot N, Luyt CE, Schmidt M, Combes A. Severe pulmonary embolism in COVID-19 patients: a call for increased awareness. Crit Care. 2020;24(1):274.

22. Artifoni M, Danic G, Gautier G, Gicquel P, Boutoille D, Raffi F, et al. Systematic assessment of venous thromboembolism in COVID 19 patients receiving thromboprophylaxis: incidence and role of D dimer as predictive factors. Journal of Thrombosis and Thrombolysis. 2020; 50(1):211–216.

23. Fraissé M, Logre E, Pajot O, Mentec H, Plantefève G, Contou D. Thrombotic and hemorrhagic events in critically ill COVID-19 patients: a French monocenter retrospective study. Crit Care. 2020;24(1):275.

24. Thomas W, Varley J, Johnston A, Symington E, Robinson M, Sheares K, et al. Thrombotic complications of patients admitted to intensive care with COVID-19 at a teaching hospital in the United Kingdom. Thromb Res. 2020; 191:76–77

25. Lodigiani C, Iapichino G, Carenzo L, Cecconi M, Ferrazzi P, Sebastian T, et al. Venous and arterial thromboembolic complications in COVID-19 patients admitted to an academic hospital in Milan, Italy. Thromb Res. 2020;191:9–14.

26. Poissy J, Goutay J, Caplan M, Parmentier E, Duburcq T, Lassalle F, et al. Pulmonary Embolism in Patients With COVID-19: Awareness of an increased prevalence. Circulation. 2020;142(2):184–186.

27. Gervaise A, Bouzad C, Peroux E, Helissey C. Acute pulmonary embolism in non-hospitalized COVID-19 patients referred to CTPA by emergency department. Eur Radiol. 2020;1–8.

28. Longchamp A, Longchamp J, Manzocchi-Besson S, Whiting L, Haller C, Jeanneret S, et al. Venous thromboembolism in critically Ill patients with COVID-19: Results of a screening study for deep vein thrombosis. Res Pract Thromb Haemost. 2020;4(5):842–7.

29. Beun, R., Kusadasi, N., Sikma, M., Westerink, J., and Huisman, A. (2020). Thromboembolic events and apparent heparin resistance in patients infected with SARS-CoV-2. doi: 10.1111/ijlh.13230.

30. Middeldorp, S. (2020). Incidence of venous thromboembolism in hospitalized patients with COVID-19. J Investig Med High Impact Case Rep. doi: 10.1177/2324709620925571 10.1111/jth.14888.

31. Tavazzi G, Civardi L, Caneva L, Mongodi S, Mojoli F. Thrombotic events in SARS-CoV-2 patients: an urgent call for ultrasound screening. Intensive Care Med. (2020) 46:1121–3. doi: 10.1007/s00134-020-06040-3

32. Leonard-Lorant I, Delabranche X, Severac F, Helms J, Pauzet C, Collange O, et al. Acute pulmonary embolism in COVID-19 patients on CT angiography and relationship to D-dimer levels. Radiology. 2020;201561.

33. Grillet F, Behr J, Calame P, Aubry S, Delabrousse E. Acute pulmonary embolism associated with COVID-19 pneumonia detected by pulmonary CT angiography. Radiology. 2020;201544.

34. Konstantinides, S. V., Meyer, G., Becattini, C., Bueno, H., Geersing, G. J., Harjola, V. P., … & Kucher, N. (2020). 2019 ESC Guidelines for the diagnosis and management of acute pulmonary embolism developed in collaboration with the European Respiratory Society (ERS) The Task Force for the diagnosis and management of acute pulmonary embolism of the European Society of Cardiology (ESC). European heart journal, 41(4), 543–603.

35. Lyhne, M. D., Kline, J. A., Nielsen-Kudsk, J. E., & Andersen, A. (2020). Pulmonary vasodilation in acute pulmonary embolism–a systematic review. Pulmonary Circulation, 10(1), 2045894019899775.

36. Patel, D., Lakhkar, A., & Wolin, M. S. (2017). Redox mechanisms influencing cGMP signaling in pulmonary vascular physiology and pathophysiology. In Pulmonary Vasculature Redox Signaling in Health and Disease (pp. 227–240). Springer, Cham.

37. Becattini, C., Manina, G., Busti, C., Gennarini, S., & Agnelli, G. (2010). Bosentan for chronic thromboembolic pulmonary hypertension: findings from a systematic review and meta-analysis. Thrombosis research, 126(1), e51–e56.

38. Kooter, A. J., IJzerman, R. G., Kamp, O., Boonstra, A. B., & Smulders, Y. M. (2010). No effect of epoprostenol on right ventricular diameter in patients with acute pulmonary embolism: a randomized controlled trial. BMC pulmonary medicine, 10(1), 18.

39. Tejero, J., Shiva, S., & Gladwin, M. T. (2019). Sources of vascular nitric oxide and reactive oxygen species and their regulation. Physiological reviews, 99(1), 311–379.

40. Kline, J. A., Puskarich, M. A., Pike, J. W., Zagorski, J., & Alves, N. J. (2020). Inhaled nitric oxide to control platelet hyper-reactivity in patients with acute submassive pulmonary embolism. Nitric Oxide, 96, 20–28.

41. Ghofrani, H. A., Humbert, M., Langleben, D., Schermuly, R., Stasch, J. P., Wilkins, M. R., & Klinger, J. R. (2017). Riociguat: mode of action and clinical development in pulmonary hypertension. Chest, 151(2), 468–480.

42. Tamargo, J., Duarte, J., Caballero, R., & Delpón, E. (2010). Cinaciguat, a soluble guanylate cyclase activator for the potential treatment of acute heart failure. Curr Opin Investig Drugs, 11(9), 1039–1047.

43. Thoonen, R., Cauwels, A., Decaluwe, K., Geschka, S., Tainsh, R. E., Delanghe, J., … & Sips, P. (2015). Cardiovascular and pharmacological implications of haem-deficient NO-unresponsive soluble guanylate cyclase knock-in mice. Nature communications, 6(1), 1–12.

